# Determinants of adolescent lung function in Indians: race, nutrition and systemic inflammation

**DOI:** 10.1101/2021.03.01.21252646

**Authors:** Mohit Aggarwal, Anubhuti Bansal, Bapu Koundinya Desiraju, Shailendra Singh, Anurag Agrawal

## Abstract

**Rationale:** While determining normal variation of spirometric volumes, the geo-political construct of ‘Indian’ inadequately captures the diverse racial structure and varied lifestyles that exist for 1 in 5 people globally. It is necessary to determine the degree of racial heterogeneity and other underlying factors to know whether lower spirometric volumes of ‘Indians’ is normal or abnormal.

**Objectives:** To investigate the differences in spirometric volumes and their associations with overall health parameters, for adolescent children and young adults, across geo-ethnic regions within India.

**Methods:** Data was analyzed for 2338 healthy subjects aged 9-19 years with acceptable spirometry from SOLID cohort. Associations between lung function and potential determinants (Race, anthropometry, nutrition, family/life history, airway oscillometric parameters and systemic inflammation) were examined by using multiple regression. Latent profile analysis (LPA) was performed for 903 subjects to uncover hidden sub-phenotypes.

**Results:** FEV_1_ and FVC varied significantly between Indian genetic ancestries, being highest in Tibeto-Burmans and lowest in Dravidians. After statistical adjustments, FEV_1_ and FVC were positively correlated with waist-height-ratio, shoulder-height-ratio, normalized BMI, and blood hemoglobin. Lower FVC was associated with lower expiratory flows (PEF and FEF_25-75_), higher lung reactance (X_5_), higher airway resistance (R_10_ and R_15_), gastro-intestinal symptoms, and higher inflammation (IL-8 and IL-17). A sub-phenotype of thinness, higher inflammation (IFN-gamma, IL-17, TNF-alpha, IL-8) and lower FVC was identified on LPA for 35% of the sample.

**Conclusions:** Spirometric volumes in Indians are different between major racial subgroups but may also be abnormally low due to prevalent nutritional or environmental adversity. Care must be taken while establishing normative standards.

## INTRODUCTION

Spirometry, the most commonly used pulmonary function test, has been through many standardization guidelines. Spirometry volumes are seen to vary widely between apparently healthy people, even after accounting for obvious anthropometric determinants like height, weight, and gender. Other than the ratio of forced expiratory volume in 1 s to forced vital capacity (FEV_1_ / FVC), where fixed cutoffs are possible even if not recommended, other parameters are absolute values that need to be interpreted against the predicted normal. This is statistically simple but physiologically complex. Differences in genetic backgrounds, environmental exposures and co-morbid conditions lead to variations that may be as large as those seen in disease. For example, South Asians have 30% lower median FEV_1_ or FVC than White Europeans or Americans (1), the lowest in the world; well within the range of pathological reduction for a White person. The solution so far has been to adjust the normal based on race, ethnicity or geography; imperfect surrogates for differences in nature and nurture (2). This creates further problems when these stereotypes are poorly thought out, as in the case of ‘Blacks’. Use of wrong normal values has associated problems of bias and justice, ranging from ineligibility for occupations with respiratory health thresholds when normal values are overestimated, to denial of medical care and disability relief when normal values are underestimated (3, 4). Here, we examine the concept of spirometric normal for ‘Indians’ – which is a geo-political construct, used as a general label for 1 in 5 people in the world, but failing to recognize genetic diversity second only to Africa and wide variations in living conditions (5). The Indian Genome Variation Database (IGVDb) has cataloged genetic variation within India and identified regions of relatively distinct genetic ancestries referred to as Indo-European (IE), Dravidian (DR) and Tibeto-Burman (TB), as well as areas of admixture (6). While there are many reports of Southern regions of India (DR ancestry) having lower lung function than Northern regions (IE and TB ancestry), this has never been tested in an equivalent nation-wide group of subjects (7). Heterogeneity of socio-economic backgrounds and environmental exposures further complicates matters (8, 9). It has been previously reported in the journal that children from rural and semi-urban India have poorer lung function than those in cities or their Indian counterparts in UK (10). Similarly, it has been shown that Indian-origin first generation Americans have better lung function than their parents, but worse than that of other Americans (11). To better understand and more finely resolve the heterogeneity of lung function in India, we studied adolescent children from a residential school system across India that caters to local populations and is relatively homogenous from a lower-middle socioeconomic class standpoint. We chose the second decade of life as our target age-group since this is when lung growth is maximal and adult lung function is reached. Also, diseases and co-morbidities other than asthma, which may affect lung function, are uncommon during this period. We focused on identifying ethnic, lifestyle and personal predictors, as well as relating spirometry measurements to lung oscillometry and other non-respiratory measures of health. We find that there is much complexity in ‘normal’ lung function of ‘Indians’ and rational non-discriminatory systems to assess general and respiratory health through spirometry are currently lacking, with ‘Indian’ or ‘South Asian’ being an inadequate but widely used label. Some of the results of these studies have been previously reported in the form of an abstract (12).

## METHODS

### Study Participants and Setting

Study participants were enrolled from 14 districts from a residential school system (Jawahar Navodaya Vidyalaya) run by an autonomous government organization (Navodaya Vidyalaya Samiti). Children in these boarding schools mostly belong to lower-middle socio-economic background and stay in a similar living arrangement. School visits were conducted to all the 14 sites and participants of Classes VI, IX, and XII (aged 9-19 years) were screened from 2017-2019. Subjects with asthma were removed from the analysis.

2609 subjects (1210 girls, 1399 boys) from 14 Indian districts were examined in SOLID cohort. Data was analyzed for 2338 subjects with acceptable spirometry with no asthma (Table 1). Oscillometry and blood investigations were performed in 1493 and 1582 subjects, respectively (Figure 1).

**Table 1:**
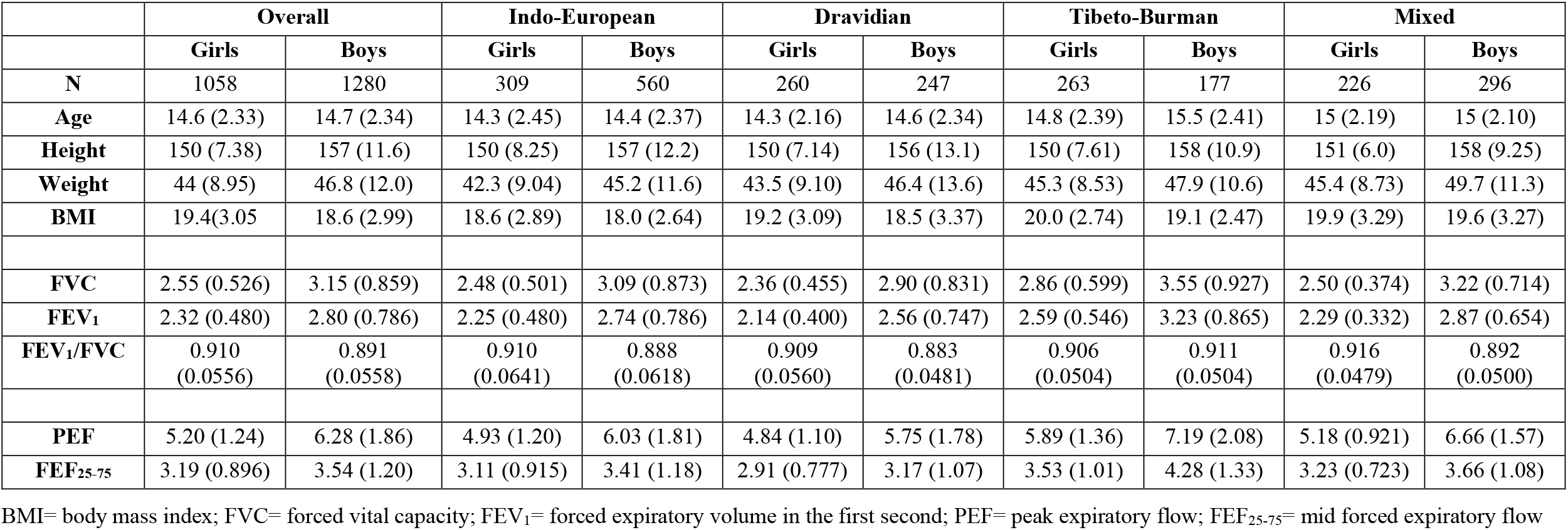
Study Participants description.

**Figure 1:**
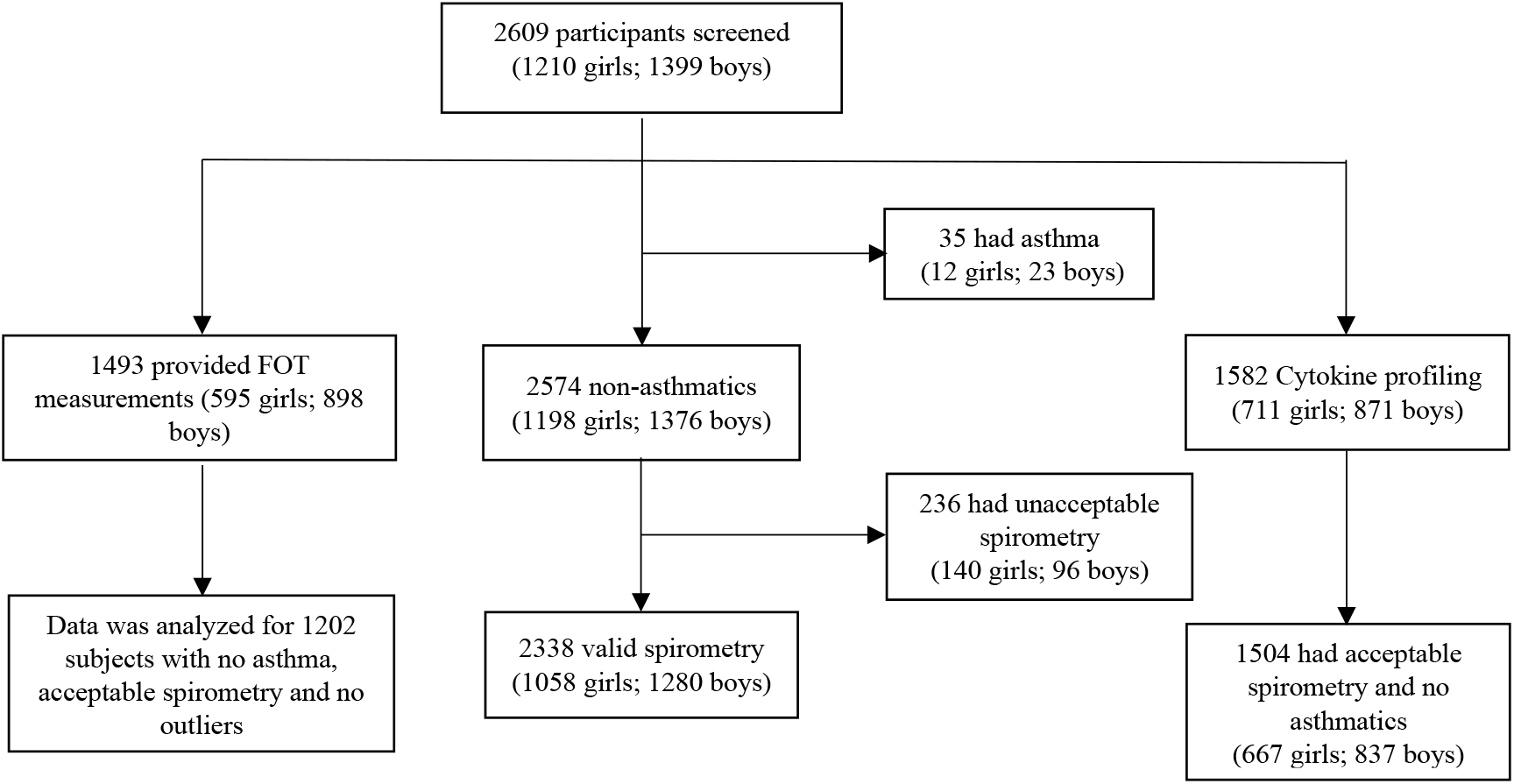
Study profile (SOLID cohort) 2609 subjects (aged 9-20 years) were screened in the SOLID cohort between 2017-2019. SOLID: Study of Lung function and Its Development, FOT: Forced Oscillation Technique

Approval of the human ethical committee of CSIR-Institute of genomics and integrative biology was obtained for the present study (Ref No: CSIR-IGIB/IHEC/2017-2018). Written Parental consent and verbal assent were taken for all the participants.

### Lung Function Assessment and its Definition

Spirometry was done using portable spirometers (EasyOne air, NDD) as per the ATS guidelines (13). Airway oscillometry was performed on 1497 SOLID participants using forced oscillation technique (Pulmoscan, Cognita Labs). A dataset from 51 older subjects who underwent oscillometry on well-established MasterIOS with acceptable spirometry was used to verify oscillometric findings from this study.

Absolute values of spirometric indices were converted to z-scores using the reference equations from GLI (Global Lung Initiative) and Chhabra et al (14, 15). Reference equations were developed internally for the PulmoScan from the SOLID data, to calculate normalized predicted values for comparisons only within this cohort (supplementary S2). R_5_ measurements were excluded from the association analysis as its reference equations had unacceptably poor fit.

### Blood Sample Collection and Biochemical Analysis

About 8 ml of whole blood was collected in vacutainer with no additives (Red top Vacutainers, BD). The Blood was allowed to clot and the supernatant was removed post centrifugation. Both the supernatant and blood pellets were stored at −80 degrees Celsius for further investigation. Cytokine levels were measured in the supernatant using a custom MILLIPLEX MAP Human Cytokine/Chemokine Magnetic Bead Panel (Merck Millipore) of 9 different cytokines. The prescribed standard assay protocol was used and the data was acquired on a magnetic bead compatible assay analyzer (Magpix, Luminex corp.). The data was analyzed using the xPONENT 3.1 software (Luminex corp.). MFI (Mean Fluorescence Intensity) values obtained were transformed to concentration (pg/mL) using a 5-parameter logarithmic standard curve. Cut-off from Mayo clinic laboratories were used to define subjects with “High” and “Normal” levels of these cytokines(16).

### Anthropometry and Other Assessments

Height, weight, shoulder width and waist circumference were measured for the anthropometry. Z-scores of BMI was calculated using the World health organization (WHO) growth chart (17). A parental and subject questionnaire was administered having questions related to background, habits and symptoms in the SOLID cohort. Genetic ancestry was mapped at the level of geographic regions that have previously been mapped by the Indian Genome Variation Database (IGVdb) to correspond to distinct or mixed genetic ancestries denoted as Indo-European (IE), Dravidian (DR), Tibeto-Burman (TB), or mixed (6). It is noted that individuals were not mapped to specific ancestries, only the geographic regions. Since geo-ethnic, linguistic and genetic maps in India are broadly concordant, this approach provides useful information in resolving the contributors to variation in lung function across India, without making assumptions about individual participants based on appearance or names.

### Statistical Analysis

Associations of lung functions were investigated using multiple (linear) regression models adjusted for covariates (age, sex, height, altitude and ethnicity). Association of each determinant was tested separately and not adjusted for the other potential lung function determinants. Cytokine levels were logarithmically transformed, using base e (2.71828), to measure their association with the lung function. A model-based clustering method (Latent profile analysis) was used to segregate 903 SOLID participants, with numerical data available for WHtR and cytokine levels, into groups based on their anthropometry and inflammatory profile. The method of LPA has already been explained extensively elsewhere(18). Interquartile (IQR) method was used to remove outliers for lung function, anthropometry and natural logarithm of cytokine levels for the cluster analysis. Values were removed if they were 1.5 times IQR below or above the first and third quartile, respectively. Pearson’s correlation coefficient was calculated to define the association amongst the model parameters and with zFVC. Listwise deletion was used to remove the missing values for a multivariate analysis. Latent profile analysis was performed using the “mclust” package (version 5.4.6) implemented in R(19). Bayesian Information Criteria (BIC) was used for the optimal model selection. An ellipsoidal model with equal orientation was selected consisting of two classes (Supplementary: Figure S4). Lung function and model parameters were compared between these two classes by independent t-test. R (version 3.6.1) was used for data analysis and visualization.

## RESULTS

Data was analyzed for 2338 (1058 girls, 1280 boys) healthy subjects with no asthma and acceptable spirometry (Figure 1). Within this group, cytokine markers were measured in 1504 subjects and oscillometry data was available for 1202 subjects. As expected, boys had higher absolute values of height, weight, FVC, FEV_1_, PEF and FEF_25-75_ and lower BMI, compared to girls (Table 1). Age and height were similar across the groups. There were modest differences in weight and BMI across regions, with IE being the lowest and TB/mixed being the highest.

### Variations in lung function based on ethnicity, habits and life-history

Table 2 summarizes the variability of lung function after adjustment for age, sex, height, altitude and ethnicity as covariates. TB group had the highest lung volumes (FVC and FEV_1_) followed by IE, after adjusting for all remaining covariates. DR group was characterized by the poorest lung function. Birth weight and non-vegetarian diet had no influence over the adolescent/adult lung volumes of the study population. Lower birth weight subjects however had slightly higher FEV_1_/FVC ratio. Subjects with family history of asthma and allergic diseases had lower FEV_1_/FVC ratio but similar lung volumes. Type of diet showed no association with the lung function, however skipping meals and chronic digestive health issues (gastrointestinal symptoms) were associated with a reduction in lung volumes.

**Table 2:** Variations in lung function based on ethnicity and life history. Linear regression was performed to calculate the association with lung function after adjusting for age, height, sex, altitude and ethnicity (for ethnicity all the remaining covariates were adjusted). Statistical significance was assessed by a two-sided p-value

|  |  | FVC |  |  | FEV<sub>1</sub> |  |  | FEV<sub>1</sub>/FVC |  |  |
| --- | --- | --- | --- | --- | --- | --- | --- | --- | --- | --- |
|  | N | Estimate | 95% CI | p-value | Estimate | 95% CI | p-value | Estimate | 95% CI | p-value |
| **Ethnicity** |  |  |  |  |  |  |  |  |  |  |
| Indo-European | 869 (37.2%) |  |  |  |  |  |  |  |  |  |
| Dravidian | 507 (21.7%) | **-0.13** | **(-0.17, -0.09)** | **<0.001** | **-0.12** | **(-0.16, -0.09)** | **<0.001** | -0.002 | (-0.008, 0.004) | 0.56 |
| Tibeto-Burman | 440 (18.8%) | **0.17** | **(0.11, 0.22)** | **<0.001** | **0.18** | **(0.13, 0.23)** | **<0.001** | 0.006 | (-0.002, 0.015) | 0.12 |
| Mixed | 522 (22.3%) | 0.01 | (-0.03, 0.05) | 0.52 | 0.02 | (-0.01, 0.06) | 0.21 | 0.004 | (-0.002, 0.01) | 0.2 |
| **Birth Weight** |  |  |  |  |  |  |  |  |  |  |
| 2.5-2.9 Kg (Average) | 1047 (50%) |  |  |  |  |  |  |  |  |  |
| <2.5 Kg (Low) | 338 (16.1%) | -0.017 | (-0.06, 0.03) | 0.45 | 0.007 | (-0.034, 0.049) | 0.72 | **0.009** | **(0.002, 0.016)** | **0.017** |
| >2.9 Kg (High) | 711 (33.9%) | -0.002 | (-0.04, 0.03) | 0.9 | -0.005 | (-0.04, 0.028) | 0.78 | -0.001 | (-0.006, 0.005) | 0.81 |
| **Diet** |  |  |  |  |  |  |  |  |  |  |
| Vegetarian/Ovo-Vegetarian | 646 (27.7%) |  |  |  |  |  |  |  |  |  |
| Non-vegetarian | 1687 (72.3%) | 0.03 | (-0.01, 0.07) | 0.2 | 0.01 | (-0.03, 0.05) | 0.61 | -0.003 | (-0.01, 0.003) | 0.26 |
| **Skip meals** |  |  |  |  |  |  |  |  |  |  |
| Never/Rarely | 1996 (85.9%) |  |  |  |  |  |  |  |  |  |
| Often/Sometimes | 327 (14.1%) | **-0.08** | **(-0.12, -0.03)** | **<0.001** | **-0.06** | **(-0.1, -0.02)** | **0.004** | 0.004 | (-0.003, 0.011) | 0.23 |
| **Digestive health problems** |  |  |  |  |  |  |  |  |  |  |
| Never/Rarely | 2141 (92.2%) |  |  |  |  |  |  |  |  |  |
| Often/Sometimes | 182 (7.8%) | **-0.08** | **(-0.13, -0.02)** | **0.008** | **-0.09** | **(-0.14, -0.03)** | **0.001** | -0.003 | (-0.012, 0.006) | 0.49 |
| **Tingling sensation or numbness in hands and feet** |  |  |  |  |  |  |  |  |  |  |
| No | 1978 (90.1%) |  |  |  |  |  |  |  |  |  |
| Yes/Maybe | 218 (9.9%) | **-0.08** | **(-0.13, -0.02)** | **0.0054** | **-0.06** | **(-0.11, -0.02)** | **0.01** | 0.002 | (-0.006, 0.009) | 0.69 |
| **Anemia** |  |  |  |  |  |  |  |  |  |  |
| Non-Anemia | 639 (31.7%) |  |  |  |  |  |  |  |  |  |
| Mild | 710 (35.3%) | 0.004 | (-0.03, 0.04) | 0.83 | -0.01 | (-0.05, 0.02) | 0.54 | -0.003 | (-0.009, 0.003) | 0.27 |
| Moderate | 652 (32.4%) | **-0.07** | **(-0.11, -0.03)** | **0.001** | **-0.07** | **(-0.11, -0.04)** | **<0.001** | -0.004 | (-0.01, 0.003) | 0.25 |
| Severe | 13 (0.6%) | -0.057 | (-0.25, 0.14) | 0.57 | -0.08 | (-0.26, 0.1) | 0.39 | -0.011 | (-0.041, 0.019) | 0.48 |
| **zBMI for age** |  |  |  |  |  |  |  |  |  |  |
| Average ($\geq 2$) | 2146 (92.9%) | | | | | | | | | |
| Low ($< -2$) | 164 (7.1%) | **-0.3** | **(-0.36, -0.24)** | **<0.001** | **-0.2** | **(-0.27, -0.17)** | **<0.001** | **0.018** | **(0.009, 0.027)** | **<0.001** |
| **Waist to Height ratio** |  |  |  |  |  |  |  |  |  |  |
| Average | 2084 (90%) |  |  |  |  |  |  |  |  |  |
| Low (below 5th percentile) | 116 (5%) | **-0.24** | **(-0.31, -0.17)** | **<0.001** | **-0.15** | **(-0.21, -0.85)** | **<0.001** | **0.02** | **(0.01, 0.03)** | **<0.001** |
| High (Above 95th percentile) | 115 (5%) | **0.13** | **(0.06, 0.19)** | **<0.001** | 0.03 | (-0.04, 0.09) | 0.42 | **-0.03** | **(-0.04, -0.02)** | **<0.001** |
| **Smokers in family** |  |  |  |  |  |  |  |  |  |  |
| No | 1725 (74.4%) |  |  |  |  |  |  |  |  |  |
| Yes | 595 (25.6%) | 0.02 | (-0.013, 0.056) | 0.229 | 0.026 | (-0.006, 0.058) | 0.11 | 0.003 | (-0.003, 0.008) | 0.3 |
| **Family history of allergic disease/atopic dermatitis/asthma** |  |  |  |  |  |  |  |  |  |  |
| No | 2046 (93.1%) |  |  |  |  |  |  |  |  |  |
| Yes | 151 (6.9%) | 0.008 | (-0.053, 0.069) | 0.8 | -0.04 | (-0.092, 0.019) | 0.2 | **-0.013** | **(-0.022, -0.004)** | **0.004** |

zBMI= z-score of body mass index for age; FVC= forced vital capacity; FEV<sub>1</sub>= forced expiratory volume in the first second, zBMI= z-score of body mass index; CI= confidence interval

### Association of lung function with markers of growth and nutrition

Waist to height ratio (WHtR), as the marker of central adiposity, showed a positive association with lung volumes after adjustment for ethnicity and other covariates (Table 3). Subjects with broader shoulders proportional to height (SHtR) and a higher z-score of BMI for age showed improved lung volumes, but a lower FEV_1_/FVC ratio. Hemoglobin levels were also positively associated with the lung volumes, with moderately severe anemia being associated with significantly reduced lung function (Table 2). There were very few subjects with severe anemia but size of effect was similar to that of moderate anemia, although not reaching statistical significance.

**Table 3:**
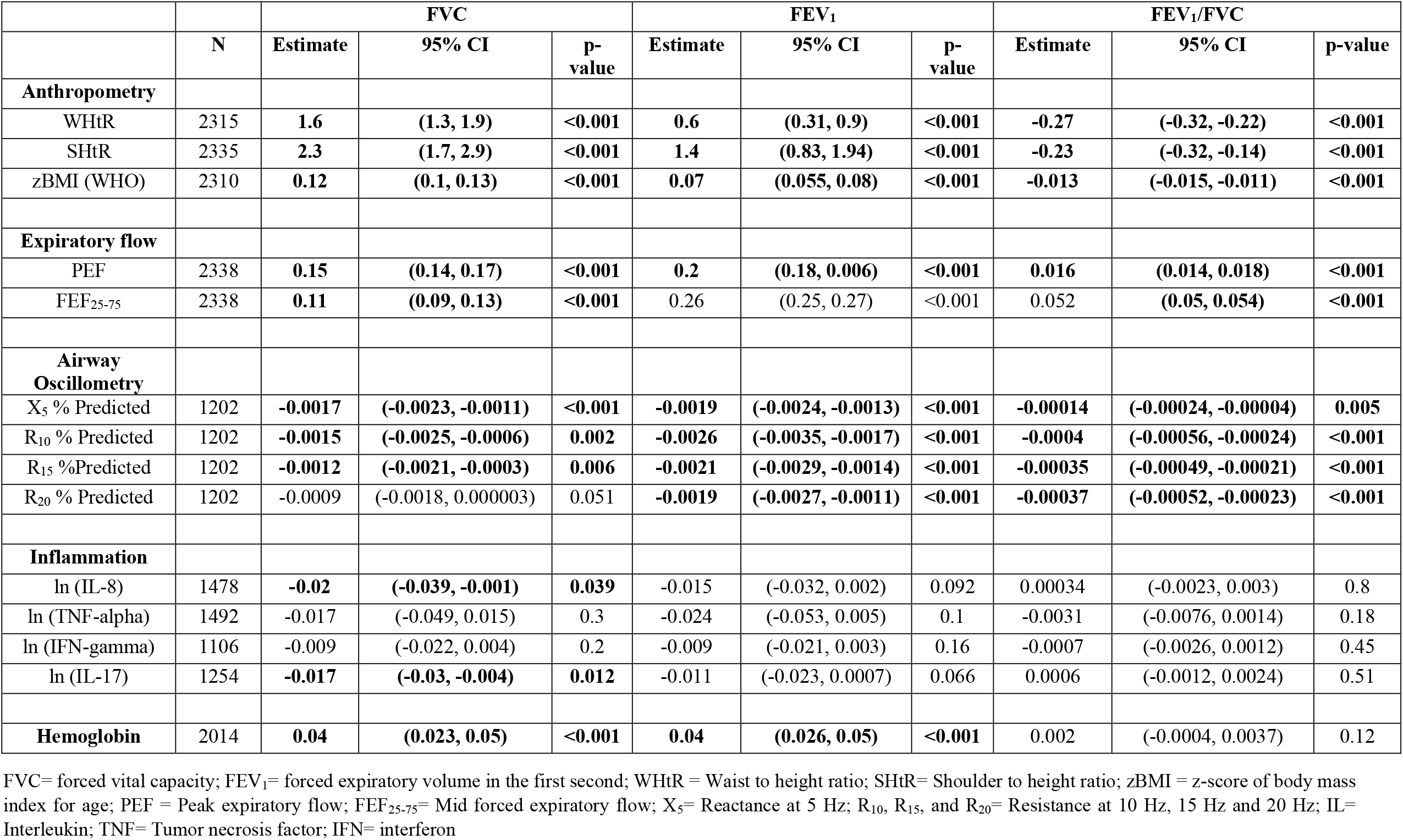
Association of lung function with growth and other health parameters. Linear regression was performed to calculate the association with lung function after adjusting for age, height, sex, altitude and ethnicity. Statistical significance was assessed by a two-sided p-value

### Association of lung function with airflow and oscillometric parameters

Peak flow (PEF) and mid-expiratory flow (FEF_25-75_) was positively associated with both the lung volumes and FEV_1_/FVC ratio (Table 3). Percent predicted values of reactance (X_5_ % predicted) calculated from internal reference equations showed a negative association with all the three lung function parameters, such that low FEV_1_ or FVC or FEV_1_/FVC ratio corresponded to higher reactance. Similarly, percent predicted values of resistance at different frequencies (10, 15 and 20 Hz) were higher in subjects with lower FEV_1_, FVC and FEV_1_/FVC ratio. Since increased reactance and resistance may correspond to sub-clinical airway pathology, we additionally verified the association of lower FEV_1_ or FVC (normal FEV_1_/FVC ratio), with markers of sub-clinical airway pathology in PFT lab data from 51 apparently healthy adult Indian subjects who underwent oscillometry (Supplementary: Figure S1).

### Systemic inflammation and lung function

In view of data suggesting potential association of lower FEV_1_ or FVC in these children with poorer general, digestive, or respiratory health, we additionally looked at associations with markers of systemic inflammation. Inflammatory studies showed a very high prevalence of clinically elevated levels of IL-8 and TNF-alpha, based on available normal ranges from western data (Supplementary: Figure S2). Many subjects also had “High” levels of IL-4, IL-17, and IFN-gamma. Prevalence of high IL-13, IL-6 and IL-1beta was very minimal. There are no specific reference values for India. IL-8 and IL-17 showed a negative association with FVC but not with FEV_1_ and FEV_1_/FVC.

### Global reference equation and latent profile analysis

Given the associations of adiposity, digestive health, nutritional markers, and inflammation, with lung function – particularly FVC – we further determined if there was a profile associated with poorer lung function in India, which operates across ethnicities. Global z-scores of forced vital capacity (zFVC) were calculated for all subjects, independent of ethnicity, from GLI equations for Caucasian ethnic group. zFVC_GLI_ (without any ethnic adjustment) also showed similar associations with central obesity and inflammatory parameters (Supplementary: Figure S3). zFVC_GLI_ was positively associated with WHtR and negatively with IL-8, TNF-alpha, IFN-gamma and IL-17. Subjects with low lung function from Caucasian equation but normal according to Indian standard showed greater inflammation, small airway abnormalities and poor growth (Supplementary: Table S1).

Latent profile analysis was performed to find sub-classes within the zFVC_GLI_ spectrum. Two potential sub-classes (C1 and C2) were identified (Table 4, Figure 2) using central obesity and inflammatory parameters. Subjects belonging to C1 were younger, had lower adiposity (WHtR), greater inflammation and more restrictive lung function. The inflammatory pattern of C1 was most strongly characterized by differences in IFN-gamma (2.6-fold elevation in mean absolute values versus C2), followed by IL-17 (1.8-fold), TNF-alpha (1.6-fold) and IL-8 (1.1-fold).

**Table 4:** Mean comparison of latent profiles. Table summarizes the mean of age, lung function (zFVC) and model parameters (WHtR, and natural logarithms of inflammatory cytokines: IL-8, TNF-alpha, IFN-gamma, IL-17). P values were calculated using independent T test.

|  | Cluster 1 | Cluster 2 | P-Value |
| --- | --- | --- | --- |
| **Total: N(%)** | 313 (34.7%) | 590 (65.3%) | - |
| Age (Years) | **14.2 (2.25)** | **15.0 (2.26)** | **<0.0001** |
| **Girls: N** | 82 | 304 | - |
| Age (Years) | **13.3 (2.27)** | **14.9 (2.22)** | **<0.0001** |
| **Boys: N** | 231 | 286 | - |
| Age (Years) | **14.6 (2.15)** | **15.0 (2.29)** | **0.03** |
| **Ethnicity** |  |  |  |
| Indo-European | 140 (44.7%) | 212 (35.9%) | - |
| Dravidian | 68 (21.7%) | 131 (22.2%) | - |
| Tibeto-Burman | 41 (13.1%) | 98 (16.6%) | - |
| Mixed | 64 (20.4%) | 149 (25.3%) | - |
| **zFVC<sub>GLI</sub>** | **-1.37 (0.880)** | **-1.20 (0.929)** | **0.008** |
| **WHtR** | **0.424 (0.0216)** | **0.456 (0.0409)** | **<0.0001** |
| **ln (IL-8)** | **2.70 (0.723)** | **2.58 (0.953)** | **0.027** |
| **ln (TNF-alpha)** | **3.26 (0.352)** | **2.79 (0.563)** | **<0.0001** |
| **ln (IFN-gamma)** | **2.30 (1.23)** | **1.35 (1.33)** | **<0.0001** |
| **ln (IL-17)** | **1.85 (1.17)** | **1.24 (1.39)** | **<0.0001** |

**Figure 2:**
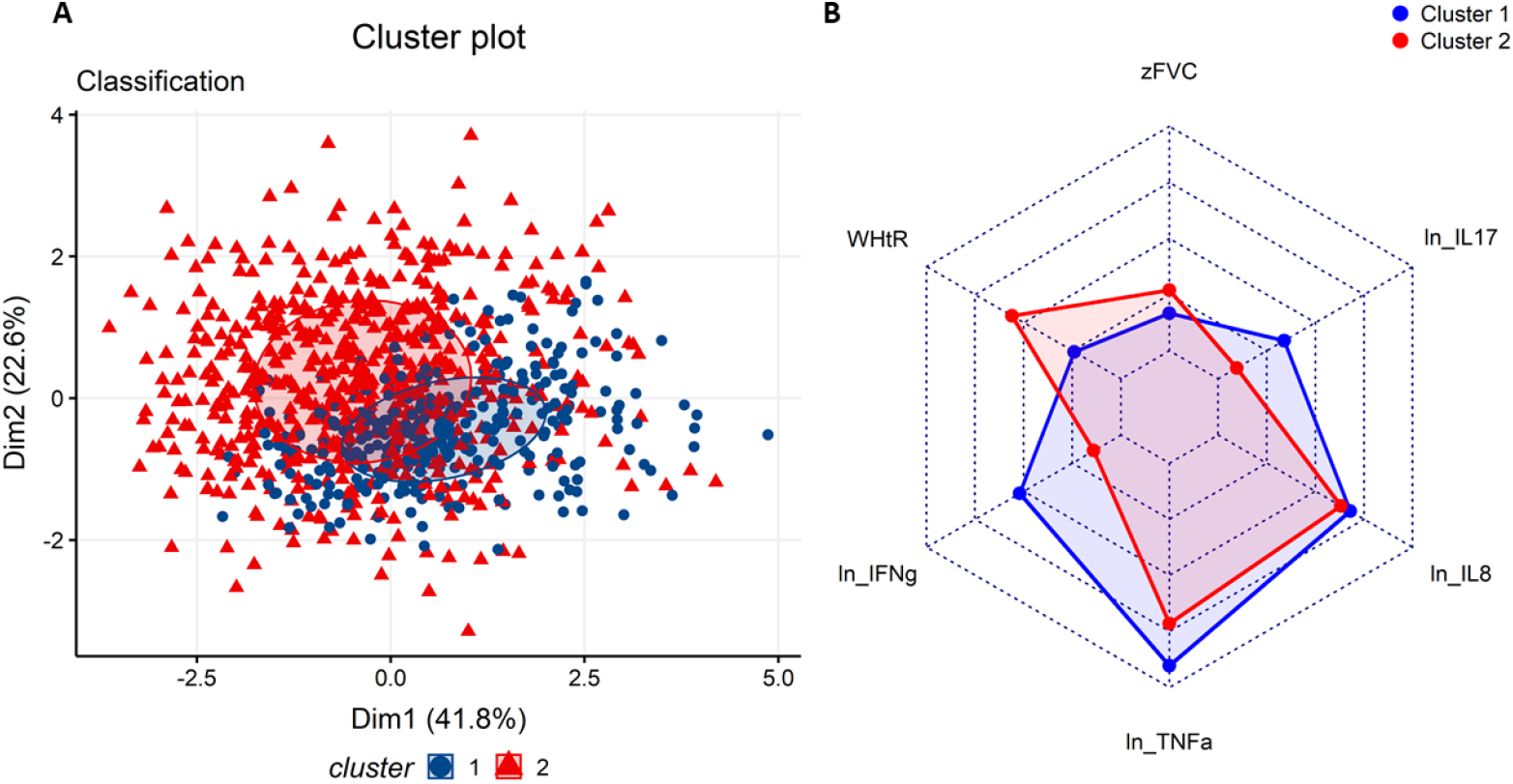
Model-based clustering using anthropometry and inflammatory parameters. (A) Scatter plot showing 2 clusters (latent profiles) generated from 903 SOLID participants using latent profile analysis: Cluster 1 (N=313) and cluster 2 (N=590). (B) Radar Plot showing the relative distribution of lung function (zFVC), anthropometry (WHtR) and inflammatory parameters (natural logarithms of IL-8, IL-17, TNF-alpha, IFN-gamma) between 2 clusters zFVC = z-scores of forced vital capacity. WHtR = Waist to height ratio. IL = interleukin. TNF = tumor necrosis factor. IFN = interferon.

## DISCUSSION

This study unequivocally demonstrates that there is no singular adjustment that can be made to determine normal lung function of Indians from global equations, nor is there a clearly definable ‘normal’ lung function that applies to all members of a geographic region known for racial diversity and extremely heterogenous living standards. To the best of our knowledge this is the first study from India that looks at adolescent to young adult spirometric lung function through a multi-prismatic lens combining anthropometry, ethno-genetics, life-history questionnaire, and general clinical markers of nutritional deficiency and inflammatory state. It is also the first study that looks at a nationally representative, but socio-economically homogenous group– members of a single government boarding school system that caters primarily to the majority lower-middle socioeconomic class. While our data does not reflect the upper socio-economic class, which is a limitation, the average Indian is well represented in this sample.

We show that there are clear racial differences in lung function, corresponding to well-known differences in genetic ancestry, with Dravidian Indians, having lower FEV_1_ and FVC after adjusting for all co-variates. This difference is most likely genetic and merits further exploration. We specifically note that the anthropometric parameters were better for this group, when compared to Indo-European group that had better lung function. While not included in the regression, this region has better air quality and higher socio-demographic indicators (SDI). We have not used regional SDI here since we selected for a similar socio-economic class. We did not introduce ambient air quality indicators for two reasons. First, air pollution exposure in Indian children of this socio-economic class is primarily through a life-history of indoor air pollution that is not quantifiable by ambient air quality measures or by simple questions. Second, it was clear that the lung function was higher in the Indo-Gangetic region of North India, including Delhi with possibly the worst air globally (not shown). Thus, ambient air quality was neither a good measure, nor a major determinant of lung function variations in our study. The Tibeto-Burman group had the best lung function. The reasons may be both genetic and environmental, since many of the regions with TB ancestry are at higher altitudes and the effects may be non-linear.

That ‘Indian’ normal lung function is different across its racial groups, was expected in view of existing global data as well as smaller normative lung function studies from different parts of India. There were two new hypothesis-generating findings in our study, however, that merit further debate and investigation. First, an association between low FVC and a phenotype of poor nutritional status and high inflammation. Second, an association between low FVC and sub-clinical airway pathology.

After adjusting for likely racial effects, we found that gastrointestinal symptoms, meal skipping, thinness, low BMI, vitamin deficiency symptoms, moderate to severe anemia, and elevated inflammation markers were all associated with lower FEV_1_ or FVC. A relatively well-defined endotype of poor lung function, low waist-height ratio and elevated IFN-gamma, IL-17, TNF-alpha and IL-8 was seen in one-third of the subjects, with membership across all ethnicities. This suggests that while there is a racial component to lower lung function in some Indians, especially those with Dravidian ancestry, a large fraction of Indians may not be reaching their genetic potential due to nutritional or environmental adversity. Since the study subjects are unlikely to be calorically limited, other possibilities to be investigated are low nutritional quality or gut dysbiosis leading to nutritional steal and enteropathy. We speculate that since the elevated cytokines are well-known mediators of gut immunity, specifically neutrophil mediated killing of microbes, this represents prevalent environmental enteropathy, a poorly recognized condition that is seen in low-middle income tropical countries and is thought to be driven by poor sanitation and recurring feco-oral exposures (20). If so, the route to better lung function for Indians may lie through better sanitation and nutrition. This fits the previous report in the journal that children from rural and semi-urban India have poorer lung function than those in cities or their Indian counterparts in UK (10). This also would explain why Indian-origin first generation Americans have better lung function than their parents (11).

In the SOLID cohort we found that low FEV_1_ or FVC, with normal FEV_1_/FVC ratio, was correlated with increased resistance and reactance. These may be signs of early small airway pathology, but these were field studies with a new portable oscillometer in an age-group not expected to have airway disease. Therefore, we decided to examine this further in older adults, using a combination of clinical grade oscillometry and nitrogen breath washout in a PFT lab setting. Lower FVC was associated with statistically and clinically significant elevations of X_5_ and R_5_-R_20_, suggesting that there may be prevalent sub-clinical small airway disease in Indians with spirometric lung function that is apparently normal. This is consistent with growing recognition of Preserved Ratio Impaired Spirometry (PRISm) as a transitional state for some population sub-groups that may foretell progression to clinical airway disease and increased all-cause mortality (21). This also fits the higher age-adjusted prevalence of COPD in India, as described recently in the Global Burden of Disease (GBD) investigator group led study of chronic respiratory diseases and their state level heterogeneity (22). Interestingly, even within India, COPD risk was highest in states with lowest socio-demographic development levels. Strong associations between low lung function and high cardio-respiratory disease risk have been reported starting from the Framingham cohort (23), which may be explained by systemic inflammation as a common link. This also highlights the potential of spirometry as a cheap precision health tool (24).

In summary, we report evidence that there is a racial sub-structure to normal spirometric lung function values in India. For those with Dravidian ancestry, normal values may be lower than for others. While India has not yet seen much genetic mixing between endogamous geographic regions, genetic measures of ancestry would be preferable to ethnic identities going forward. We also report that at least one-third of adolescents and young adults are likely to be not reaching their genetic potential due to adverse nutrition and environment.

## CONCLUSION

Spirometric volumes in Indians are different between major racial subgroups but may also be abnormally low due to prevalent nutritional or environmental adversity. Based on our data, respiratory disease would be underdiagnosed or underestimated in Tibeto-Burmans, but over-diagnosed or overestimated in Dravidians by a common yardstick. Care must be taken while establishing normative standards for ‘Indians’ and studies including deep phenotyping and genetic markers are required to dissect this further.

## Data Availability

Data can be provided on request on contacting the corresponding author. The data could not be provide publicly since it contain extensive individual level information that could compromise anonymity.

## Notes

### Competing Interest Statement

The authors have declared no competing interest.

### Funding Statement

1. Wellcome Trust DBT India Alliance IA/CPHS/ 14/1/501489
2. Indo-US S&T Forum grant USISTEF/HI-011/2015-16
3. CSIR MLP 5502

### Author Declarations

Approval of the human ethical committee of CSIR-Institute of genomics and integrative biology was obtained for the present study (Ref No: CSIR-IGIB/IHEC/2017-2018).

