## Supplementary for "Determinants of adolescent lung function in Indians: race, nutrition and systemic inflammation"

**Table of Contents**

**Supplementary Methods**

S1 Study population

S2 Lung function measurements

S3 Blood collection and cytokine profiling

S4 Anthropometry and other measurements

S5 Statistical analysis

**Supplementary Results**

Table S1: Comparison of means between groups with globally low but locally normal lung function and normal by both standards

Figure S1 Association of zFVC and airway oscillometry parameters in Delhi-NCR adults

Figure S2 Prevalence of clinically elevated levels of inflammatory cytokines.

Figure S3 Correlation Matrix for lung function, anthropometry and inflammation.

Figure S4 Optimal model selection using BIC.

**Supplementary References**

### METHODS

#### S1: Study Population

The study was performed in a cohort of residential school children, named SOLID (Study Of Lung function & Its Development), comprising of subjects aged 9-19 years. 14 Sites were selected across India based on diversity in geography, ethnicity and culture. 14 Districts were: Chandigarh, Jhajjar, Bharatpur, Jaisalmer, Bengaluru, Thiruvananthapuram, Puducherry, Puri, 24 North Parganas, Mandi, Ribhoi, Po-Phodong, Leh and Gumla. Study sites were selected in a manner that it would account for most of the ethnic and geographical diversity of India. A list of desirable states was provided to the Navodaya Vidyalaya Samiti (NVS) and they approved 14 schools in 11 different states and 2 union territories. Written parent consent was obtained in all the schools before the visit and subject assent was taken before the assessment. Participants with poor spirometry and asthma were excluded from the analysis.

Additional data of 51 adults from Delhi-NCR was analyzed to evaluate the associations of lung function and FOT (Forced Oscillation Technique) parameters.

#### S2: Lung Function Measurements

Spirometric measurements were performed using a portable handheld spirometer (EasyOne, NDD, Switzerland) using the ATS guidelines (1). Data for only those subjects were considered for the analysis which had at least 3 acceptable maneuvers and 2 repeatable measurements (acceptable QC scores: A, B and C). Age of subjects were calculated from the date of their birth to the date of test.

Airway oscillometry was performed using the Impulse oscillometry (MS-IOS, Jaeger) in Delhi-NCR adults and Forced oscillation technique (PulmoScan, Cognita labs, USA) in SOLID cohort. Oscillometry was performed in 8 districts of SOLID cohort: Chandigarh, Jhajjar, Bharatpur, Jaisalmer, Bengaluru, Thiruvananthapuram, Puri and 24 North Pargana. Data obtained on PulmoScan in the SOLID cohort was used for the generation of reference equations and for the calculation of predicted values of R5 (Adj. $R^2$: Girls = 0.008, Boys = 0.159), R10 (Adj. $R^2$: Girls = 0.081, Boys = 0.343), R15 (Adj. $R^2$: Girls = 0.104, Boys = 0.367), R20 (Adj. $R^2$: Girls = 0.116, Boys = 0.349) and X5 (Adj. $R^2$: Female = 0.120, Male = 0.302). Multiple regression was performed in 1179 subjects (437 girls, 742 boys) with no asthma, acceptable spirometry, $z(FEV_1/FVC) > -1.64$ and outliers removed for R5, R10, R15, R20, and X5 using standing height, age and sex as the predictors. R5 values were not included in the association analysis since its reference equations had poor values of adjusted $R^2$ (measure of goodness of fit).

**S3: Blood collection and cytokine profiling**

Blood was collected in the SOLID cohort using the plain red top vacutainers (BD). Around 8 ml blood was withdrawn from the subjects who gave their consent by pricking them only once. Serum was separated on site by centrifugation post coagulation and stored immediately in dry ice. Collected samples were transported to CSIR-IGIB, Delhi in dry ice where they were eventually stored in -80 degrees Celsius freezer. Blood samples with any red tinge were discarded on the account of hemolysis.

A custom MILLIPLEX MAP Human Cytokine/Chemokine Magnetic Bead Panel (Merck Millipore) were used to measure the levels of 9 cytokine: IL-4, IL-6, IL-8, IL-13, IL-17, IL-1 Beta, IFN-gamma, TNF-alpha and TGF-alpha. The standard assay protocol was used and a magnetic bead compatible assay analyzer (Magpix, Luminex corp.) was used for the acquisition of data. Data was analyzed using xPONENT 3.1 software (Luminex corp.). A 5-parameter logarithmic standard curve was used to transform raw MFI (Mean Fluorescence Intensity) into concentration values (pg/mL). Data points with bead count below 50 were discarded to ensure the quality of observed values. Reference values from Mayo clinic laboratories were used for the classification of subjects into groups with “High” and “Normal” levels of inflammatory cytokines(2). Data points which were below the detection limit were assigned the “Normal” label.

**S4: Anthropometry and other measurements**

A digital weighing scale (Omron) and stadiometer (IS Indosurgicals) were used for the measurement of weight and height, respectively. A parental and subject questionnaire was also administered in the SOLID cohort having questions related to symptoms, background and habits.

Genetic ancestry was mapped at the level of geographic regions that have previously been mapped by the Indian Genome Variation Database (IGVdb) (3) to correspond to distinct or mixed genetic ancestries denoted as Indo-European (IE), Dravidian (DR), Tibbeto-Burman (TB), or mixed. It is noted that individuals were not mapped to specific ancestries, only the geographic regions. Since geo-ethnic, linguistic and genetic maps in India are broadly concordant, this approach provides useful information in resolving the contributors to variation in lung function across India, without making assumptions about individual participants based on appearance or names.

**S5: Statistical Analysis**

R version 3.6.1 was used for the statistical analysis and graphical representation.

Associations of lung functions were investigated using multiple (linear) regression models adjusted for covariates (age, sex, height, altitude and ethnicity). Association of each determinant was tested separately and not adjusted for the other potential lung function determinants. Pearson’s correlation was performed to evaluate the association between continuous variables such as, zFVC, age, Oscillometric parameters and concentration of inflammatory cytokines. The null hypothesis was that there is no correlation and rejected when P value was below 0.05. Results of correlation was represented as R (Correlation coefficient), CI (95 % confidence Interval) and P value. Independent T-test was performed for the comparison of mean between two groups with null hypothesis (rejected at $P < 0.05$) that there was no difference between them.

Latent profile analysis (LPA) was performed for the identification of clusters based on Inflammatory and anthropometric parameters. It is a model-based clustering approach which tries to assume the distribution (gaussian) of hidden groups (latent profiles) based on continuous variables. Mclust package was used to perform the LPA(4). In this study, LPA was performed using the WHtR as the anthropometric variable along with Log (IL-8), Log (TNF-alpha), Log (IFN-gamma) and Log (IL-17) as the inflammatory variables. Modelling was performed for 903 subjects who had numerical responses for all these 5 variables available after the removal of outliers. Optimal model was selected by considering the highest BIC (Bayesian Information Criterion) value. A 2 class VVE (ellipsoidal, equal orientation) model was selected having the highest BIC (Fig. S4).

### RESULTS

| | GLI Low\|Indian Normal | Normal GLI & Indian | p-value |
|---|---|---|---|
| WHtR | **0.44** | **0.46** | **<0.001** |
| SHtR | **0.226** | **0.232** | **<0.001** |
| zBMI (WHO) | **-0.83** | **-0.18** | **<0.001** |
| zFEF2575 | **-0.58** | **0.0008** | **<0.001** |
| PEF | **5.2** | **6.1** | **<0.001** |
| X5 % Predicted | **107.8** | **94.9** | **<0.001** |
| R10 % Predicted | **102.7** | **98.3** | **0.001** |
| R15 % Predicted | **102.9** | **98.4** | **0.002** |
| R20 % Predicted | **102.5** | **98.6** | **0.006** |
| Hemoglobin | 11.4 | 11.5 | 0.11 |
| ln (IL-8) | **2.7** | **2.5** | **0.005** |
| ln (TNF-alpha) | **2.9** | **2.8** | **0.045** |
| ln (IFN-gamma) | 1.8 | 1.6 | 0.074 |
| ln (IL-17) | 1.33 | 1.26 | 0.45 |

**Table S1: Comparison of means between groups with globally low but locally normal lung function and normal by both standards**

GLI=Global lung Initiative, WHtR= waist to height ratio, SHtR= Shoulder to height ratio, zBMI = z-score of body mass index, $zFEF_{25-75}$= z-score of mid forced expiratory flow, PEF= peak expiratory flow, $X_5$= Reactance at 5 Hz; $R_{10}$, $R_{15}$, and $R_{20}$= Resistance at 10 Hz, 15 Hz and 20 Hz; IL= Interleukin; TNF= Tumor necrosis factor; IFN= interferon

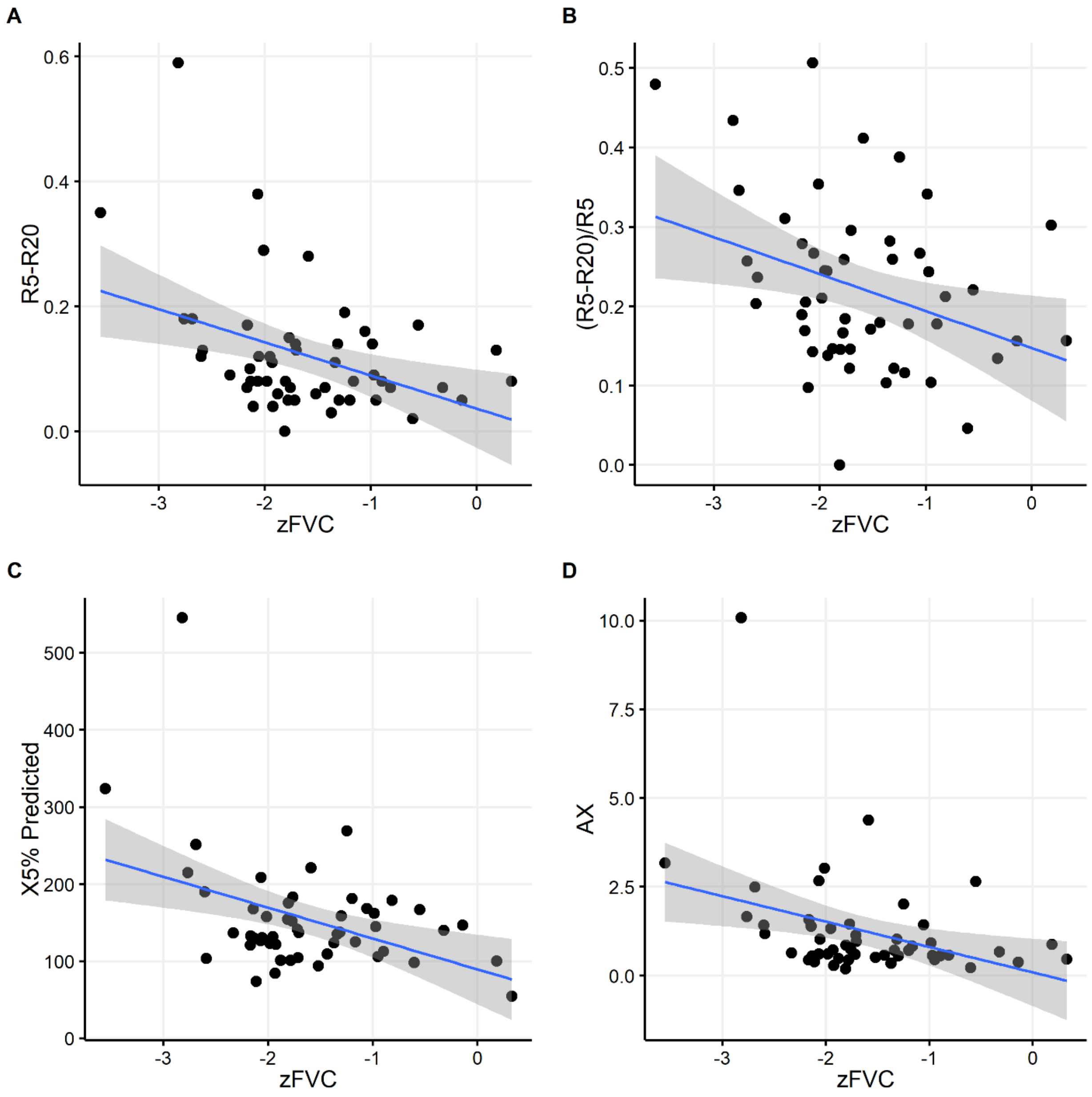

**Figure S1: Association of zFVC and airway oscillometry parameters in Delhi-NCR adults.**

Scatter plot with a linear regression curve showing association between zFVC and oscillometric parameters in 51 subjects from Delhi-NCR.

zFVC = z-scores of forced vital capacity. R5: Respiratory resistance at 5 Hz. R20: Respiratory resistance at 20 Hz. X5: Respiratory reactance at 5 Hz. AX: Area of Reactance.

| **Parameter** | **N** | **R** | **CI** | **p-value** |
|---|---|---|---|---|
| **X5% predicted** | 51 | **-0.4137** | -0.6188, -0.1559 | **0.0025** |
| **AX** | 51 | **-0.36** | -0.5782, -0.0937 | **0.0095** |
| **R5-R20** | 51 | **-0.3983** | -0.6072, -0.1378 | **0.0038** |
| **(R5-R20)/R20** | 51 | **-0.339** | -0.562, -0.07 | **0.015** |

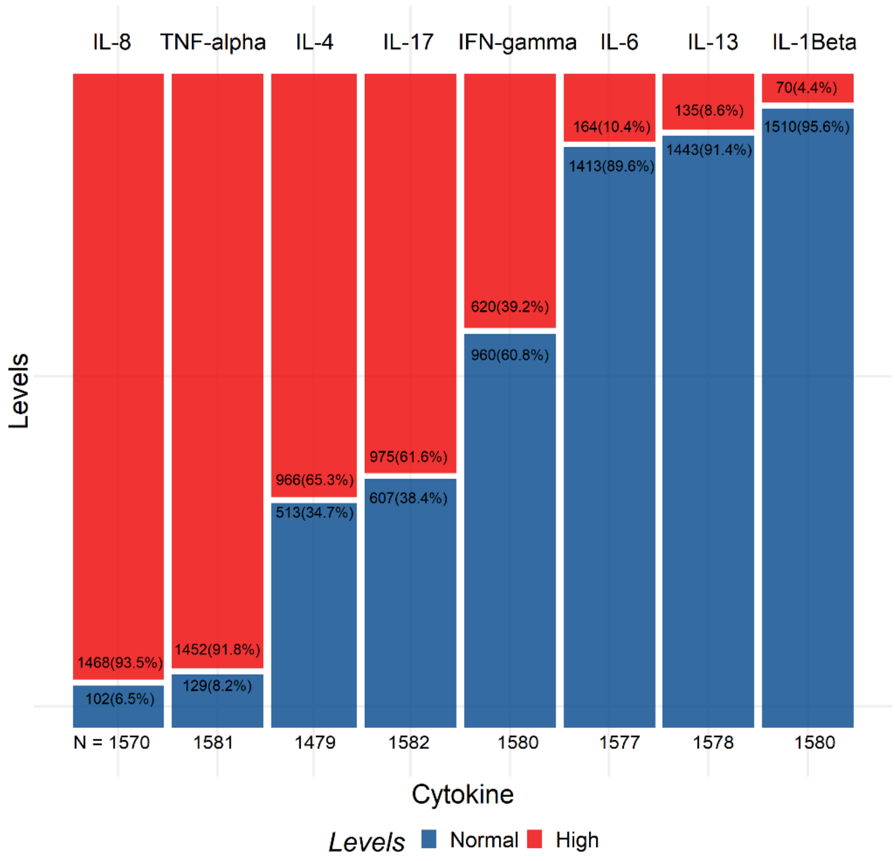

**Figure S2: Prevalence of clinically elevated levels of inflammatory cytokines.**

Mosaic plot showing the proportion of subjects with ‘High’ and ‘Normal’ levels of cytokines based on reference values from Mayo clinic laboratories.
IL = interleukin. TNF = tumor necrosis factor. IFN = interferon.

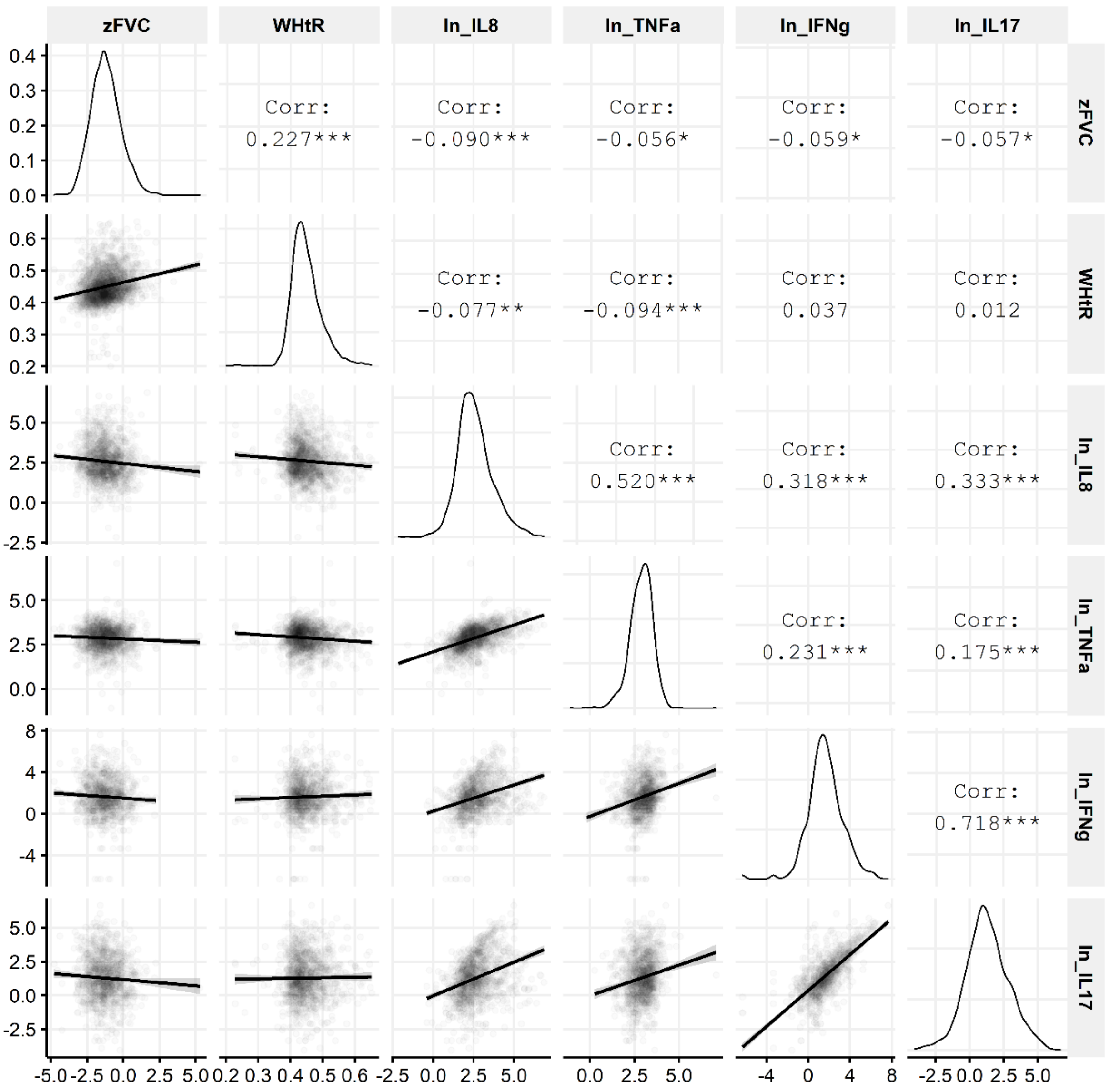

**Figure S3: Correlation Matrix for lung function, anthropometry and inflammation.**

Pearson's correlation coefficients are summarized for association amongst lung function (zFVC), anthropometry (WHtR) and natural logarithms of inflammatory cytokines: IL-8, TNF-alpha, IFN-gamma and IL-17.
zFVC: z-score of forced vital capacity, WHtR: Waist to height ratio, IL: Interleukin, TNF: Tumor necrosis factor, IFN: Interferon

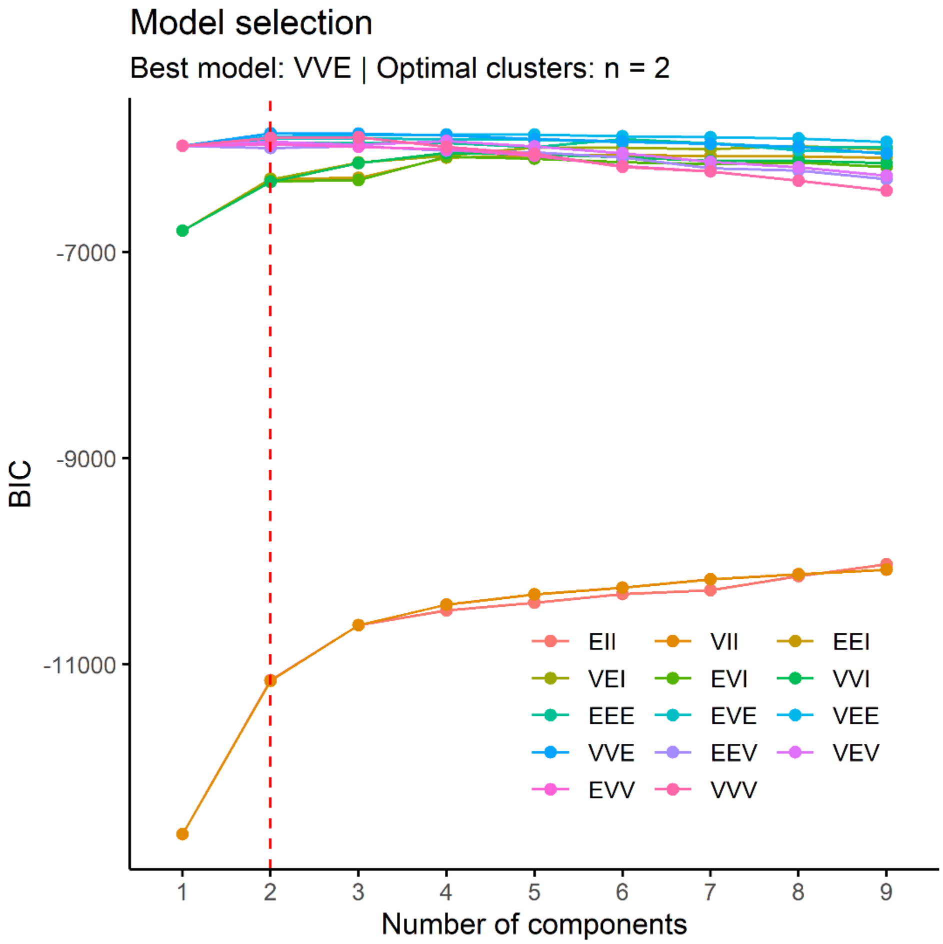

**Figure S4: Optimal model selection using BIC.**

y-axis represent the BIC values for the corresponding model with certain no of components (latent profiles) shown on the x-axis. BIC= Bayesian information criteria

A 2 component VVE (ellipsoidal, equal orientation) model was chosen based on highest BIC (-5848.356). Model was built using 5 continuous variables which showed associations with zFVC: WHtR, ln (IL-8), ln (TNF-alpha), ln (IFN-gamma), ln (IL-17).
